# CARDIO-QVARK Diagnose Ischemic Myocardiocyte!

**DOI:** 10.1101/2024.07.16.24310485

**Authors:** Basheer Abdullah Marzoog, Peter Chomakhidze, Alexander Suvorov, Philipp Kopylov

**Author notes:** Corresponding author: Basheer Abdullah Marzoog, M.D., Ph.D. cardiology student at Sechenov First Moscow State Medical University at the World-Class Research Center “Digital biodesign and personalized healthcare”. Address: Russia, Moscow, 8-2 Trubetskaya street, 119991. Postal address: Russia, Moscow, 8-2 Trubetskaya street, 119991. Scopus ID: 57486338800. Competing interests: No competing interests regarding the publication.

## Abstract

**Background:** Ischemic heart disease (IHD) has the highest mortality rate in the globe. This returns to the poor diagnostic and therapeutic strategies including the early prevention methods.

**Aims:** To assess the changes in the single channel electrocardiography (SCECG) at rest and on exercise test in patients with vs without IHD confirmed by stress computed tomography myocardial perfusion (CTP) imaging with vasodilatation stress-test.

**Objectives:** IHD frequently have preventable risk factors and causes that lead to the disease appearance. However, the lack of the proper diagnostic and prevention tools remains a global challenge in or era despite the current scientific advances.

**Material and methods:** A single center observational study included 80 participants from Moscow. The participants aged ≥ 40 years and given a written consent to participate in the study. Both groups, G1=31 with vs G2=49 without post stress induced myocardial perfusion defect, passed cardiologist consultation, anthropometric measurements, blood pressure and pulse rate, echocardiography, cardio-ankle vascular index, performing bicycle ergometry, recording 3-minutes SCECG (using CARDO-QVARK) before and just after bicycle ergometry, and then performing CTP. The LASSO regression with nested cross-validation was used to find association between CARDO-QVARK parameters and the existence of the perfusion defect. Statistical processing carried out using the R programming language v4.2, Python v.3.10 [^R], and Statistica 12 programme.

**Results:** The CARDO-QVARK parameters analysis have a specificity 75.5 % [95 % confidence interval (CI); 0.628; 0.88], sensitivity 51.6 % [95 % CI; 0.333; 0.695], area under the curve (AUC) 67 % [95 % CI; 0.530; 0.801] in compare to bicycle ergometry AUC; 50.7 % [95 % CI; 0.388; 0.625], specificity 53.1 % [95 % CI; 0.392; 0.673], sensitivity 48.4 % [95 % CI; 0.306; 0.657], based on our study results.

**Conclusion:** The SCECG have no statistically significant higher diagnostic accuracy in compare to bicycle ergometry. However, CARDO-QVARK has the potential to improve the diagnostic accuracy of the bicycle ergometry.

**Other:** Further investigations required to uncover the hidden capabilities of CARDO-QVARK in the diagnosis of ischemic heart disease.

## Introduction

Ischemic heart disease remains the leading challenge in terms of mortality and morbidity despite the advances in the used methods for diagnosis and prevention. However, the early prevention in terms of evaluation of the ischemic heart disease in early period still underestimated. The current attention of the scientists paid to the prevention rather than diagnosis and treatment. In this manner, the scientific community developed several cost-effective methods to be confirmed for clinical use for early prevention of ischemic heart disease, including the use of the single channel electrocardiography and exhaled breath analysis in coronary heart disease prevention [1].

Ischemic heart diagnosis using single channel electrocardiography (ECG) remains in the development stage and require further elaboration in the context of the sensitivity and specificity. Several kinds of single channel ECG has been used in the clinical trials including CARDO-QVARK, Apple Watch, Kardia, Zio, BioHarness, Bittium Faros and Carnation Ambulatory Monitor [1–8]. Single channel ECG has been used to diagnosis myocardial infarction and monitoring patients with chronic heart disease and heart failure as well as to classify heartbeat [9–11].

Currently there are several kinds of single channel ECG used for commercial purposes and clinical trials. The accuracy and quality of these single channel ECGs various. The uses of single channel ECGs are various including distant monitoring of patients with arrythmias, as a Holter monitoring, and for monitoring for chronic heart failure [12,13,22,14–21]. The currently available single channel ECGs in the market include Apple Watch, Kardia, Zio, Cardiostat, BioHarness, Bittium Faros and Carnation Ambulatory Monitor [13,23].

Single-channel ECG has key features that can aid in diagnosing ischemic heart disease include detecting ischemia through ECG alterations, hemodynamic changes, and clinical signs and symptoms [24]. Additionally, vectorcardiography, a technique that records cardiac electrical activity as closed loops, can be useful for training in electrocardiography and detecting cardiac ischemia [24,25]. Portable and fast electrode placement devices allow for good-quality ECG tracings, making single-channel ECG accessible and efficient [24].

In comparison of single-channel ECG to multi-channel ECG in detecting ischemic heart disease shows that modern ECG systems with vector-based electrocardiography can improve the detection of ECG alterations typical for ischemia compared to the conventional 12-lead ECG [26]. Single-channel consumer ECG devices, such as smartwatches, can be useful for detecting and monitoring arrhythmias but have limitations in detecting ST-segment deviations indicating myocardial infarction or ischemic episodes [27].

The usage of single channel in ischemic heart disease has not been previously investigated and requires further elucidation.

## Material and methods

A prospective single center cohort study included 80 participants. According to the results of the CTP, the participates divided in to two groups. The first group of participants with stress-induced myocardial perfusion defect (n=31) and the second group without stress induced myocardial perfusion defect (n = 49) in the CTP. Participants are randomly chosen. Written consent has been taken from the participants. The study registered on clinicaltrials.gov (NCT06181799), and the study approved by the ethical commitment of the Sechenov University, Russia, from “Ethics Committee Requirement № 19-23 from 26.10.2023”.

The study evaluated continuous and categorical variables. The continuous variables included; age, pulse at rest, systolic blood pressure (SBP) at rest, diastolic blood pressure (DBP) at rest, body weight, height, maximum heart rate (HR) on physical stress test, watt (WT) on physical stress test, metabolic equivalent (METs) on physical stress test, reached percent on physical stress test, ejection fraction (EF %) on echocardiography, estimated vessel age, right cardio-ankle vascular index (R-CAVI), left Cardio-ankle vascular index (L-CAVI), mean CAVI (=(right-CAVI + left-CAVI)/2), right ankle-brachial index (RABI), left ankle-brachial index (LABI), mean ankle-brachial index (ABI), mean SBP brachial (SBPB) (=(right SBPB+ left SBPB)/2), mean DBPB (=(right DBPB + left DBPB)/2), BP right brachial (BPRB) (=(SBP+DBP)/2), BP left brachial (BPLB) (=(SBP+DBP)/2), mean BPB (=(BPRB+BPLB)/2), BP right ankle (BPRA) (=(SBP+DBP)/2), BP left ankle (BPLA) (=(SBP+DBP)/2), mean BPA (=(BPRA+ BPLA) /2), right brachial pulse (RTb), left brachial pulse (LTb), mean Tb (=(LTb+ RTb)/2), right brachial-ankle pulse (Tba), left brachial-ankle pulse (Tba), mean Tba (= (left Tba+right Tba)/2), length heart-ankle (Lha in cm), heart-ankle pulse wave velocity (haPWV = Lha/(mean left Tba+ mean right Tba); m/s), β-stiffness index from PWV (=2*1050*( haPWV)^2*LN((mean SBPB *133,32/ mean DBPB *133,32))/((mean SBPB*133,32)-( mean DBPB *133,32)), creatinine (µmol/L), and eGFR (2021 CKD-EPI Creatinine). Categorical variables included; gender, obesity stage, smoking, concomitant disease, coronary artery, hemodynamically significant (>60%), myocardial perfusion defect after stress ATP, myocardial perfusion defect before stress ATP, atherosclerosis in other arteries (Yes/No), carotid atherosclerosis, brachiocephalic atherosclerosis, arterial hypertension (AH), stage of the AH, degree of the AH, risk of cardiovascular disease (CVD), stable coronary artery disease (SCAD), functional class (FC) by Watt and by METs, reaction type to stress test(positive/negative), reason of discontinuation of the stress test, CAVI degree, and ABI degree.

The study used the following SCECG parameters:

- QTc duration (Bazett’s formula);
- amplitude parameters (JA is the amplitude at point J in microV, TA is the amplitude of the T-wave in microV, PAn is the amplitude of the negative P-wave in microV);
- indices of asymmetry SBeta, Beta (ratio of the maximum modulus of the derivative value at the leading front of the T-wave to the maximum modulus of the value at the trailing front of the T-wave);
- spectral integrals of energy of R and T waves: QRS11energy (leading front of R—1st derivative), QRS12energy (trailing front of R—1st derivative), QRS2energy (R-wave as a whole—2nd derivative), TE1 (T-wave as a whole) - (the integral is calculated as the sum of energies at all points of the corresponding region);
- spectral integral set by the frequency grid 2 to 4 Hz, 4 to 8 Hz (QRSE1, QRSE2);
- frequency of the maximum energy of the leading and trailing fronts of the R wave (RonsF, RoffsF);
- rhythm variability (SDNN);
- ECG time markers: PpeakN, Rpeak, Speak, Tpeak, Tons, Toffs.

ECG time intervals were calculated not from the beginning of the cardiac cycle, but from a point on the isoline, two-thirds of the duration of the mean R-R interval from the previous R-wave (so called—Calculated point). All parameters, except for the indicators of rhythm variability, were averaged taking into account the pulse rate. Rhythm variability indicators were evaluated by the program without averaging the values. Thus, the time parameters of the ECG took into account not only the morphology of the cardiac cycle, but also changes the heart rate. Considering that averaged values were taken into account, a longer recording provides the most accurate parameters. This includes markers of the beginning or the end of the wave (Pfi, QRSst, QRSfi), the shift of the negative or positive maximum value relative to the beginning of the averaged complex (PpeakN, Rpeak, Speak, Tpeak), as well as the maximum slope of the waves (Tons, Toffs). QRSfi is the time interval from the calculated point to the end of the QRS complex, expressed in ms. Tpeak is the time interval from the calculated point to the peak of the T wave. Toffs is the time interval from the calculated point to the point of maximum steepness of the descending knee of the T wave [28].

Taking into account that the position of the calculated point depends on the R-R interval, it is possible to minimize the effect of heart rate on the time parameters of the cardiac cycle. These time parameters take into account not only the morphology of the QRS complex or T-wave, but also the temporal features of the entire cardiac cycle.

After logistic regression analysis including more than 200 ECG parameter listed above, the artificial intelligence method was used to find combinations with the highest accuracy for IHD determination.

## Criteria for the study participants

The inclusion criteria included;

1. Participants age ≥ 40 years.
2. Participants with intact mental and physical activity.
3. Written consent to participate in the study, take blood samples, and anonymously publish the results of the study.
4. The participants of the experimental group are individuals with coronary artery disease, confirmed by myocardial perfusion defect on the adenosine triphosphate stress myocardial perfusion computed tomography, and confirmed by medical history, previous medical tests, and retrospective interview of participants.

Exclusion criteria:

1. Poor single-channel ECG and pulse wave recording quality
2. Failure of the stress test for reasons unrelated to heart disease
3. Reluctance to continue participating in the study.

Non-inclusion criteria

1. Pregnancy and breast feeding.
2. Diabetes mellitus
3. Presence of signs of acute coronary syndrome (myocardial infarction in the last two days), history of myocardial infarction;
4. Active infectious and non-infectious inflammatory diseases in the exacerbation phase;
5. Respiratory diseases (bronchial asthma, chronic bronchitis, cystic fibrosis);
6. Acute thromboembolism of pulmonary artery branches;
7. Aortic dissection;
8. Critical heart defects;
9. Active oncopathology;
10. Decompensation phase of acute heart failure and mild reduced and reduced heart failure ejection fraction;
11. Neurological pathology (Parkinson’s disease, multiple sclerosis, acute psychosis, Guillain-Barré syndrome);
12. Cardiac arrhythmias that do not allow exercise ECG testing (Wolff-Parkinson-White syndrome, Sick sinus syndrome, AV block of II-III-degree, persistent ventricular tachycardia);
13. Diseases of the musculoskeletal system that prevent passing a stress test (bicycle ergometry);
14. Allergic reaction to iodine and/or adenosine triphosphate.

## Data collection

All participants at rest pass registration of S-ECG and pulse wave before (during 3 minutes) and just after (during 3 minutes) physical stress test (bicycle ergometry) using a portable single-channel recorder (CARDO-QVARK; Russia, Moscow) [29]. The SCECG and pulse wave results interpreted using machine learning models developed by the Sechenov University team [29,30].

Both groups pass a vessel stiffness test and pulse wave recording as well as vascular age by using Fukuda Denshi device (VaSera VS-1500; Japan). Cuffs placed to assess the vascular stiffness (CAVI parameter) and the vascular age as well as the ancle-brachial index [31].

Subsequently, participants pass exercise bicycle ergometry test (SCHILLER CS200 device; Bruce protocol or modified Bruce protocol). According to the results metabolic equivalent; Mets-ВT (ВТ), the angina functional class (FC) in participants with positive stress test results determined. During the bicycle ergometry, the participants monitored with 12-lead ECG and manual blood pressure measurement, 1 time each 2 minutes. The rest time ECG and blood pressure monitoring continue for at least five minutes after the end of the stress bicycle ergometry test. The positive criteria for ergometry test are the presence of ST segment depression ≥ 1 mm in any lead as well as the presence of clinical presentation of shortness of breath and typical anginal pain that disappear after discontinuing the physical test <5 minutes and or response to nitroglycerin spry inhalation.

The procedure discontinues if: an increase in systolic blood pressure ≥ 220 mmHg, horizontal or down sloping ST segment depression on the ECG ≥ 1 mm, typical heart pain during test, ventricular tachycardia or atrial fibrillation, or other significant heart rhythm disorders were found. Moreover, stop the procedure if the target heart rate (86% of the 220-age) is reached.

Before performing CTP, all the participants present results of the venous creatinine level, eGFR (estimated glomerular filtration rate) according to the 2021 CKD-EPI Creatinine > 30 ml/min/1,73 m2, according to the recommendation for using this formula by the National kidney foundation and the American Society of Nephrology [32–35].

The participants of both groups got catheterization in the basilar vein or the radial vein for injection of contrast and ATP to performed pharmacological stress test to the heart by increasing heart rate during the stressed myocardial perfusion computer tomography imaging. Computer tomography was performed on Canon scanner with 640 slice, 0,5 mm thickness of slice, with contrast (Omnipaque, 50 ml), injected two times: in rest to get images for myocardial perfusion before test, and in 20 mints just after ATP had been injected in dose according to body weight.

The results of the myocardial perfusion considered positive if there was a perfusion defect after stress test or worsen the already existing at the rest phase perfusion defect. this registry data used for development cohort, machine learning for ischemia detecting using single channel ECG.

### Statistical analysis

For quantitative parameters, the nature of the distribution (using the Shapiro-Wilk test), the mean, the standard deviation, the median, the interquartile, the 95% confidence interval, the minimum and maximum values were determined. For categorical and qualitative features, the proportion and absolute number of values were determined.

Comparative analysis for normally distributed quantitative traits was carried out on the basis of Welch’s t-test (2 groups); for abnormally distributed quantitative traits, using the Mann-Whitney U-test (2 groups).

Comparative analysis of categorical and qualitative features was carried out using the Pearson X-square criterion, in case of its inapplicability, using the exact Fisher test.

For Single channel ECG values, pre-load values (prefixed with “*q*0_” were used, and deltas between immediately after exertion (*q*1) and after 2nd single channel ECG record, were calculated:

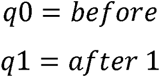

Calculation of deltas:

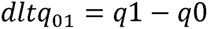

Statistical processing carried out using the R programming language v4.2, Python v.3.10 [^R], and Statistica 12 programme. (StatSoft, Inc. (2014). STATISTICA (data analysis software system), version 12. www.statsoft.com.). P considered statistically significant at <0.05.

### Outcome and feature selection

Due to the small number of observations (n = 80), random sampling of 2/3 of the available sample for predictor selection was performed for 1000 repetitions to evaluate the performance of the predictors [36]. Data preprocessing at each iteration involved normalization and iterative imputation using Bayesian ridge regression for quantitative data. There were no categorical or binary features. At each iteration, a classifier was built using the gradient boosting algorithm, which made it possible to calculate feature importances for 1000 times. Then, feature importances medians were calculated for each factor, and predictors were ranked from the highest median values to the lowest.

Ten selected predictors were included in a new pipeline, the same data preprocessing was performed, then a classifier was built using the gradient boosting algorithm. Leave-one-out cross-validation was used. After that, the area under the curve, AUC, was calculated, and the optimal threshold was selected for calculating sensitivity and specificity, positive and negative prognostic values. The obtained area under the curve was compared with the result of stress test using the McNemar criterion. This procedure was performed separately CARDO-QVARK data.

## Results

The descriptive characteristics of the sample were shown as both groups and then each group separately in tables for a full representation of the results. The characteristics of the continuous variables of the sample described in the tables below. (*Table 1A-B)*

**Table 1A:**
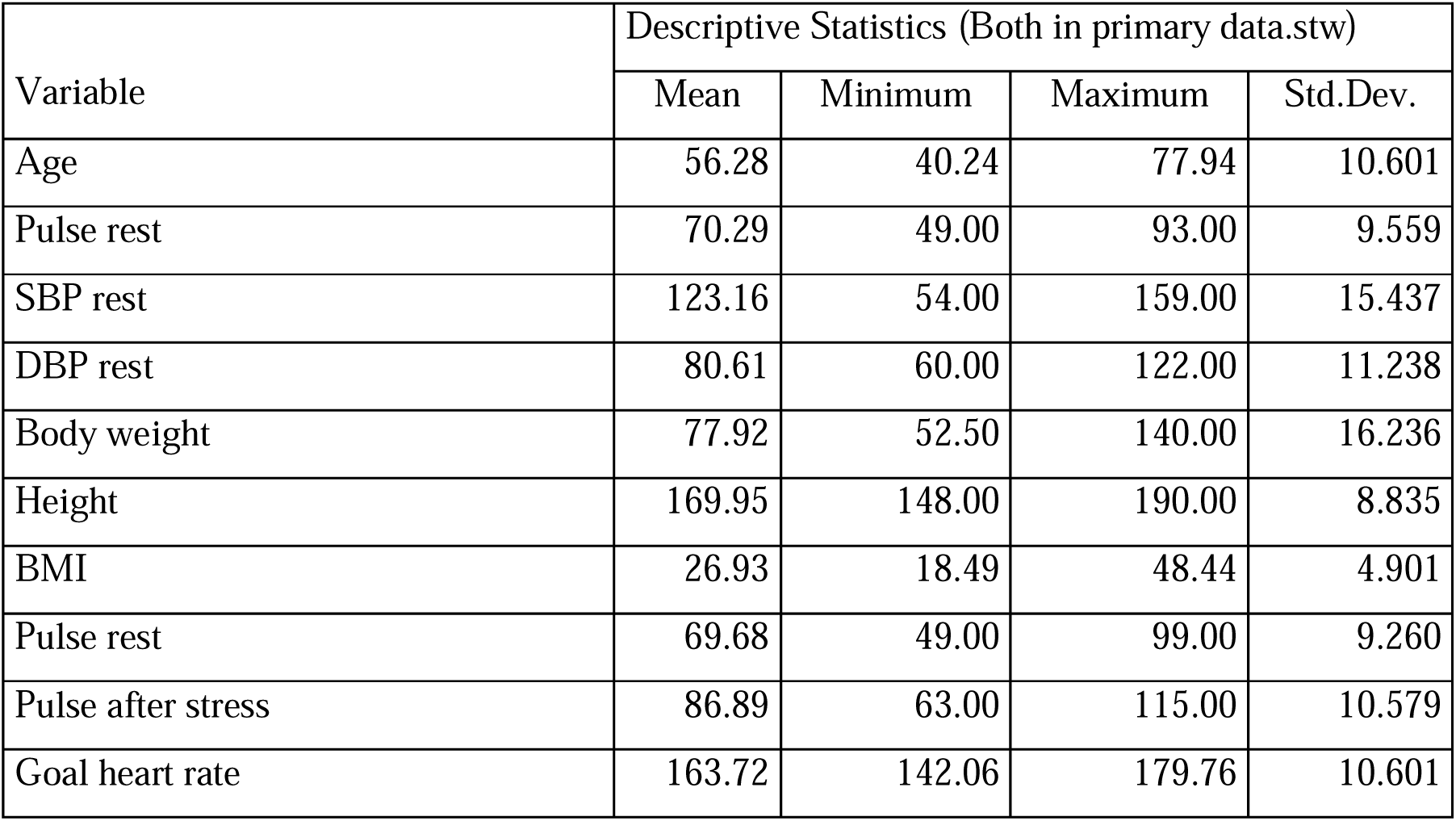
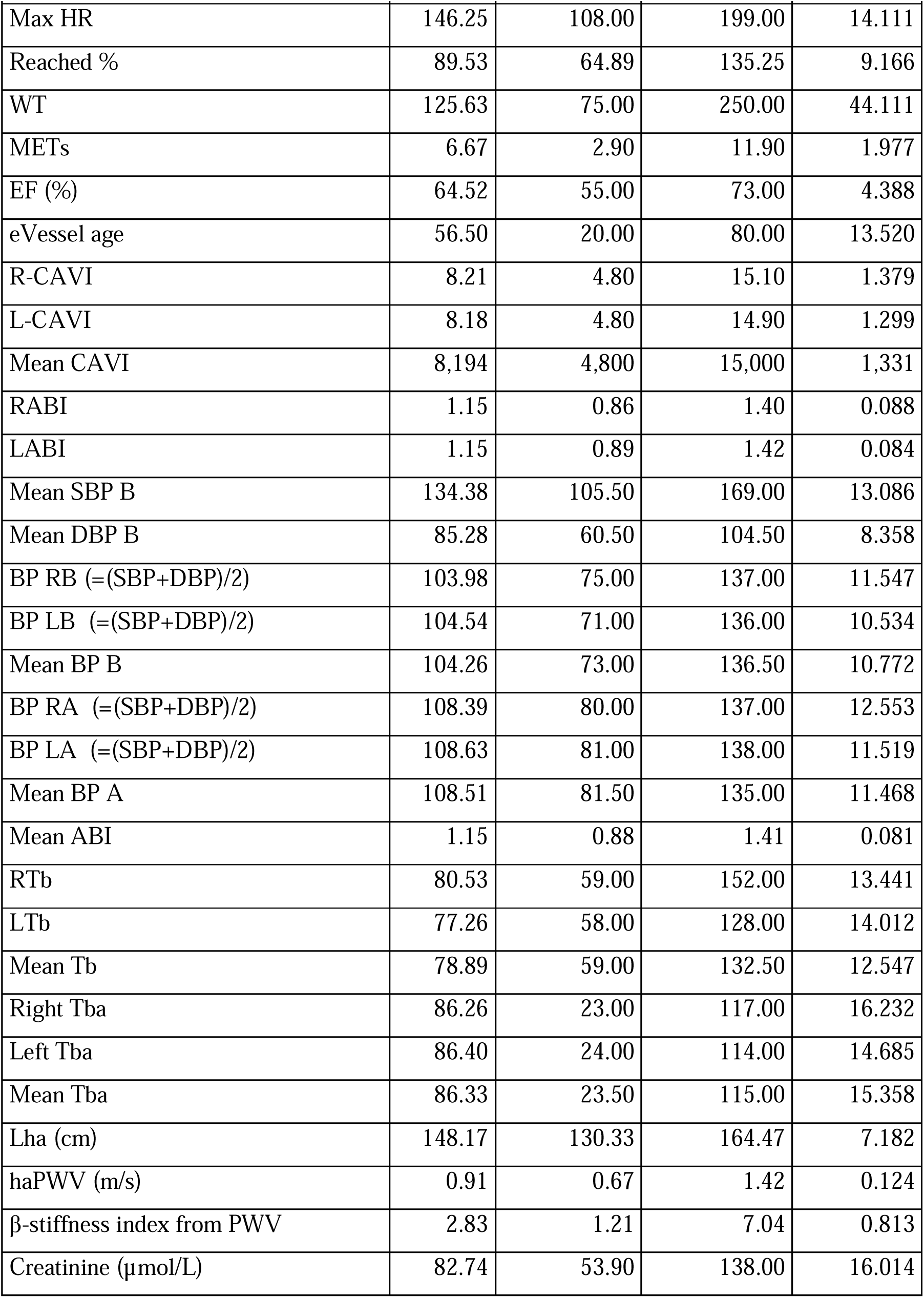
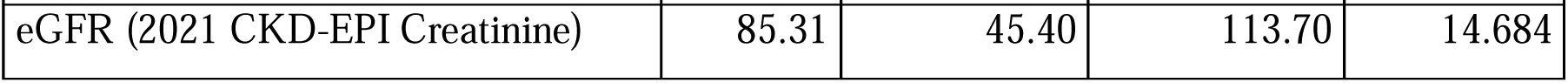
The features of the continues variables of the sample represented in the table.

**Table 1B:**
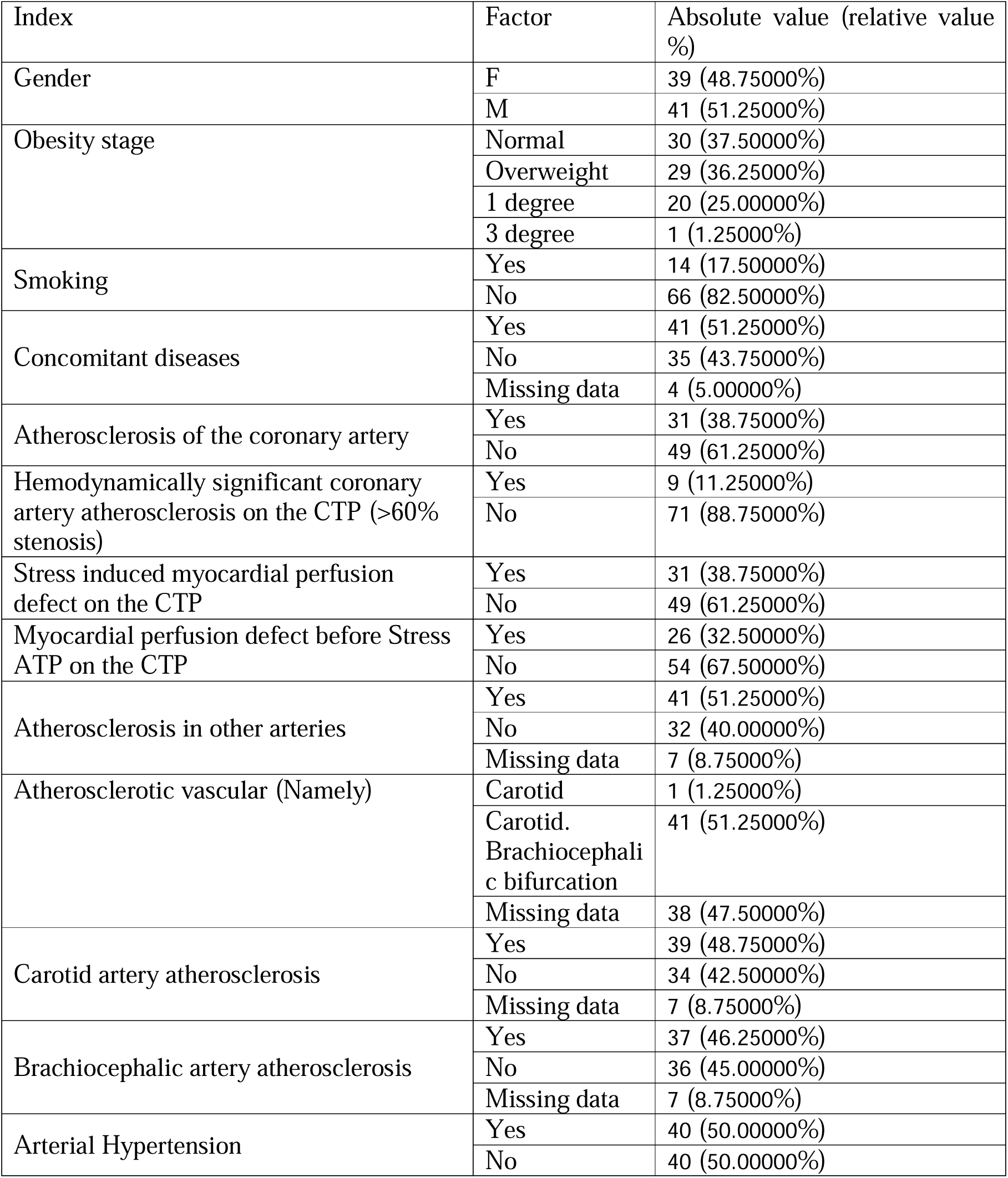
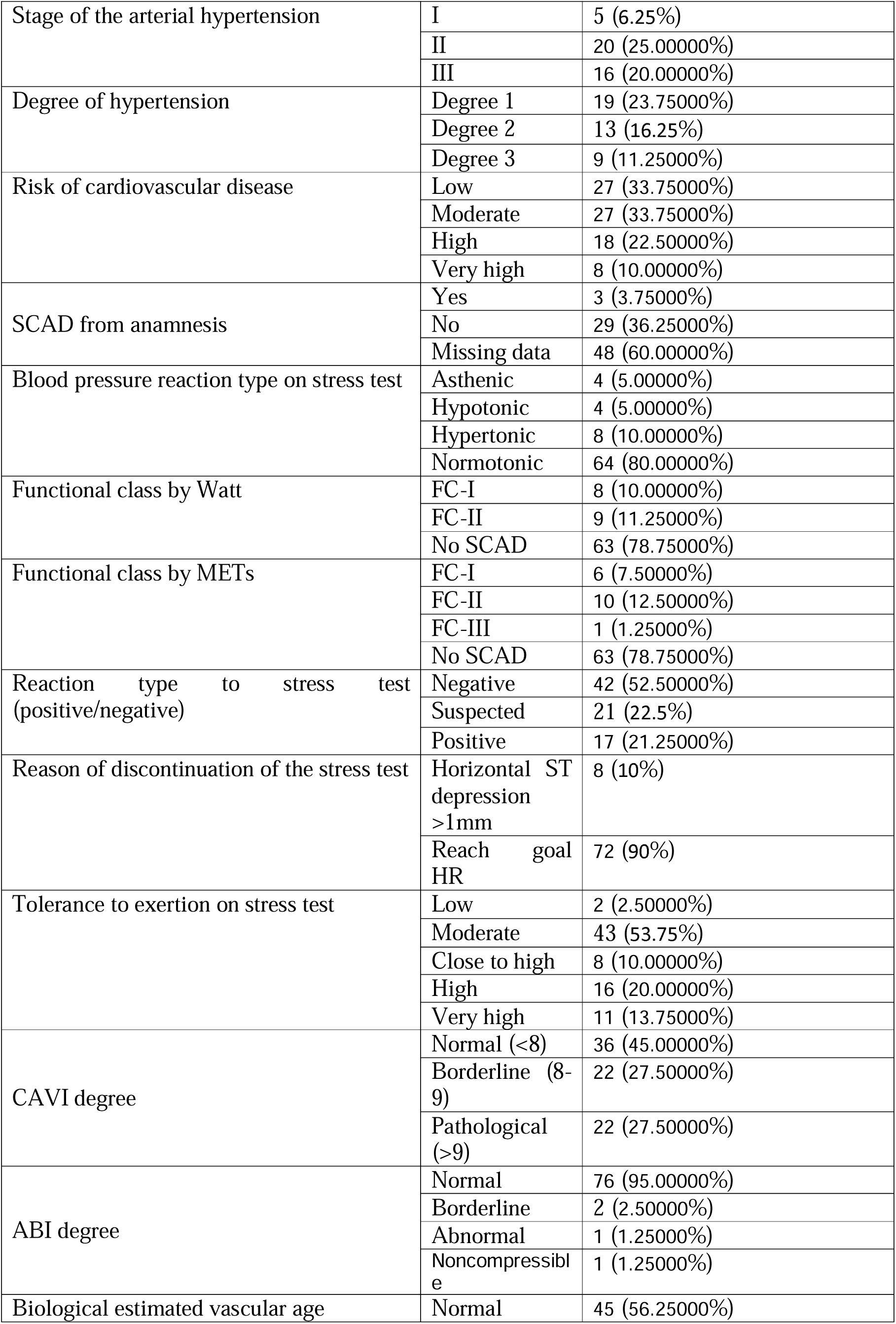
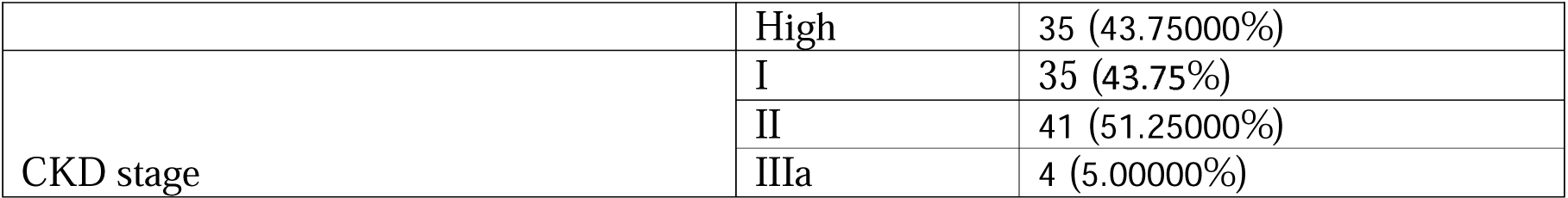
The features of the categorical variables of the sample represented in the table. Abbreviations: METs; metabolic equivalent. CPT; stress myocardial perfusion computer tomography imaging.

The comparative characteristics of the sample represented in the below tables based on the presence or absence of the stress induced myocardial perfusion defect of the CTP imaging with the adenosine triphosphate. (*Table 2A-B*)

**Table 2A:**
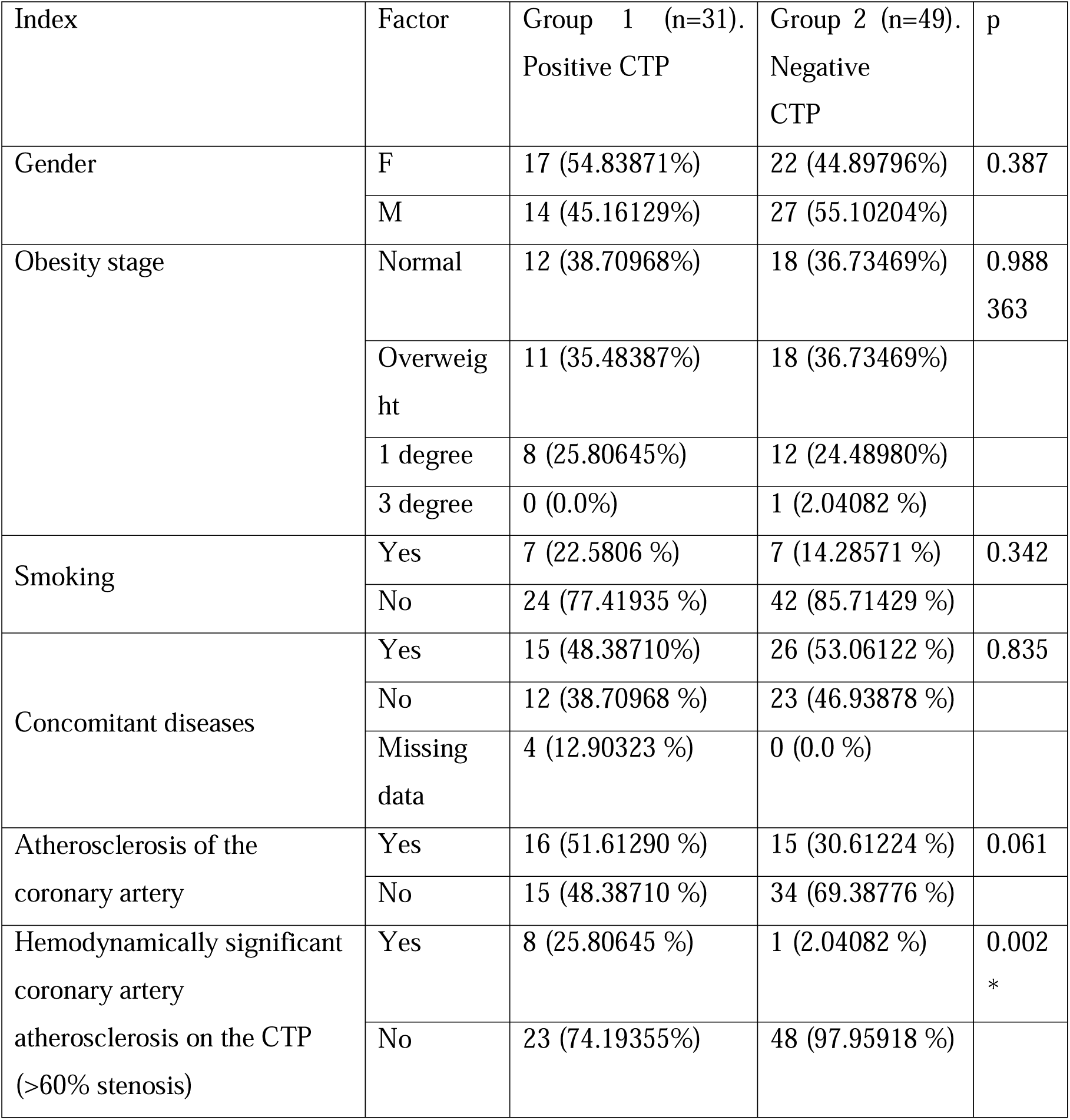
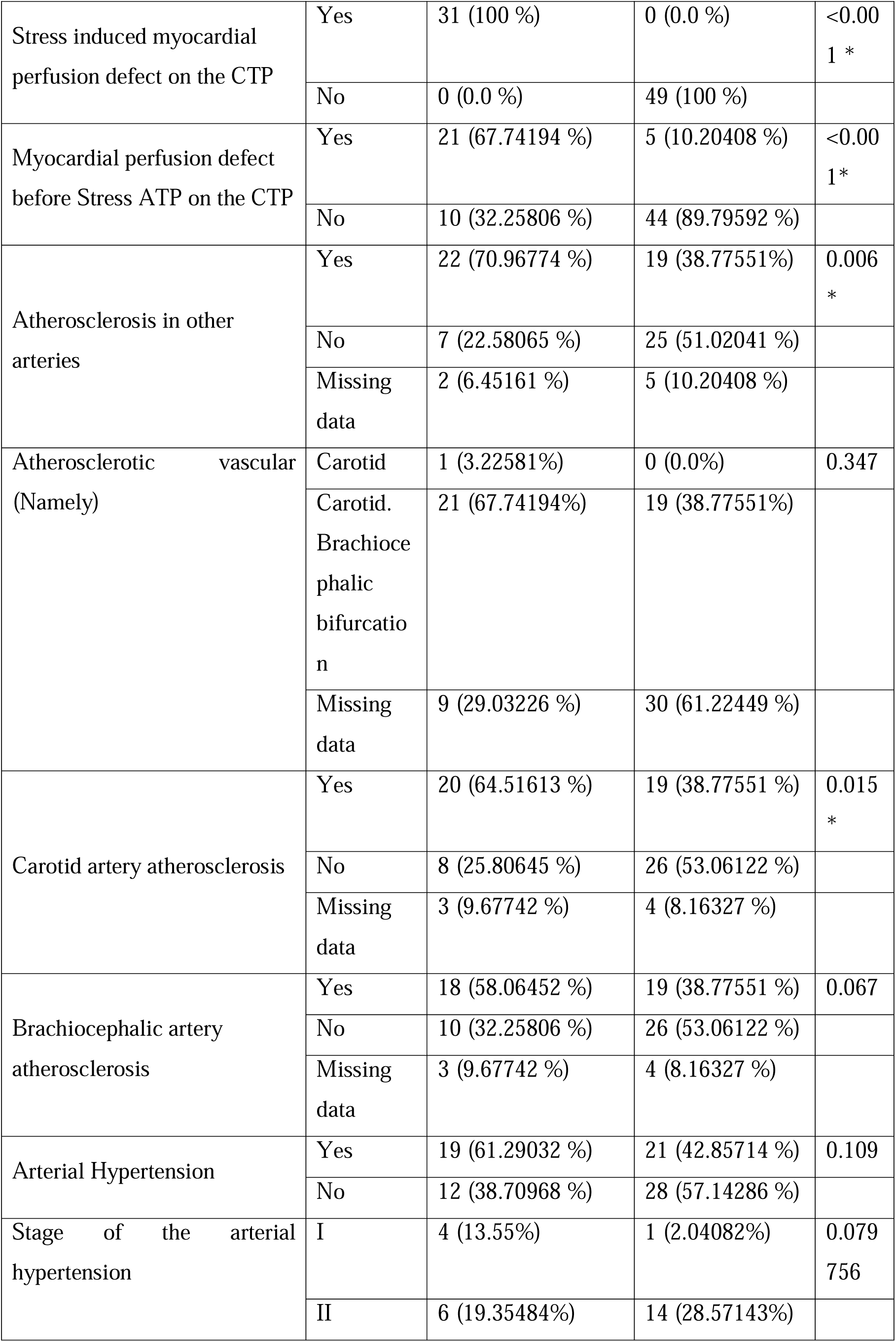
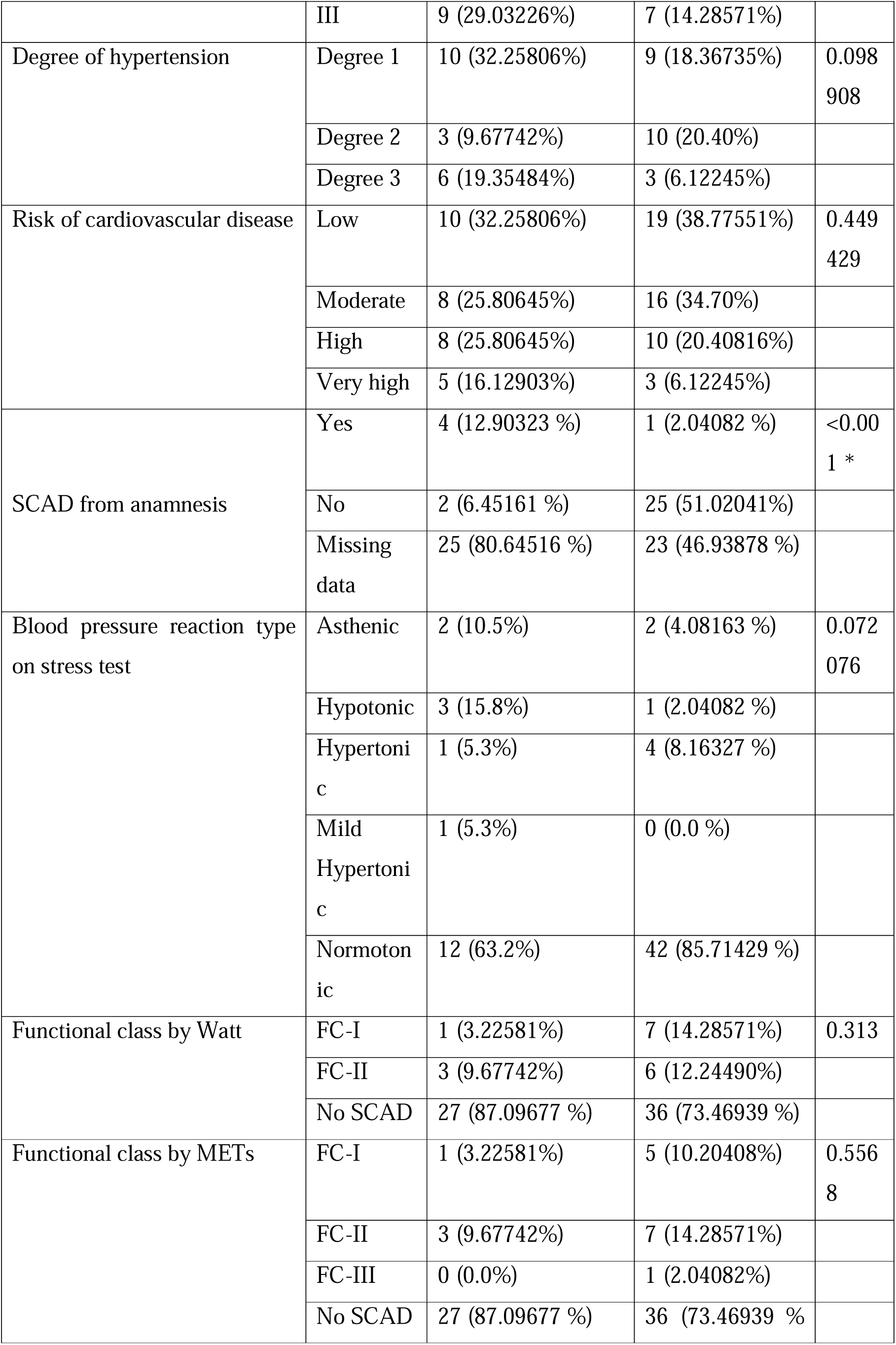
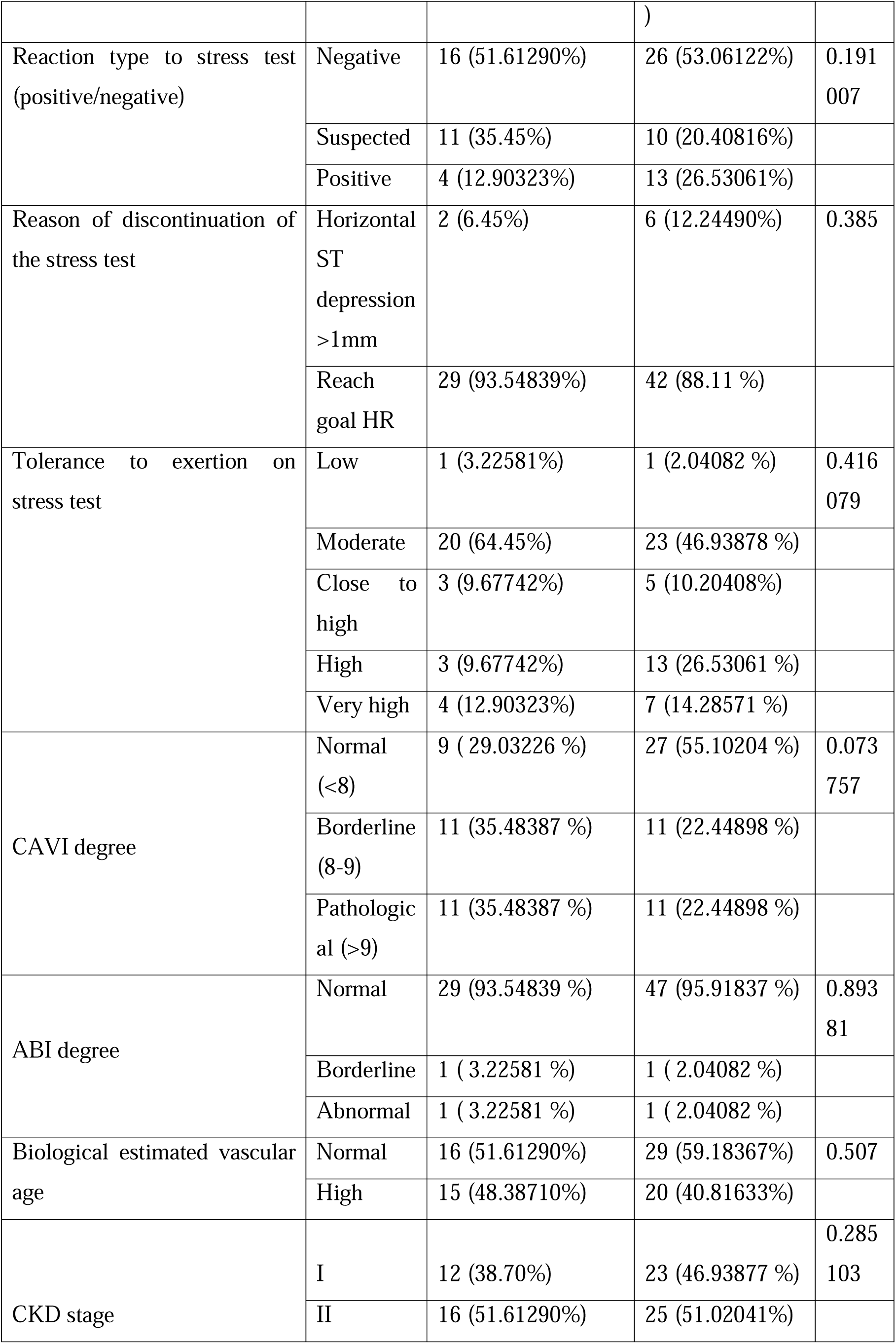
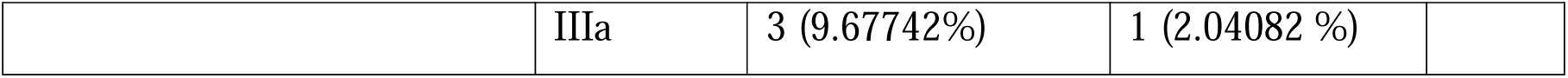
Categorical variables presented in absolute and relative study values for the true incidence of the stated factor. X^2^ test used for the analysis of categorical variables. * Values of statically significant differences. Abbreviations METs; metabolic equivalent, CPT; stress myocardial perfusion computer tomography imaging.

**Table 2B:**
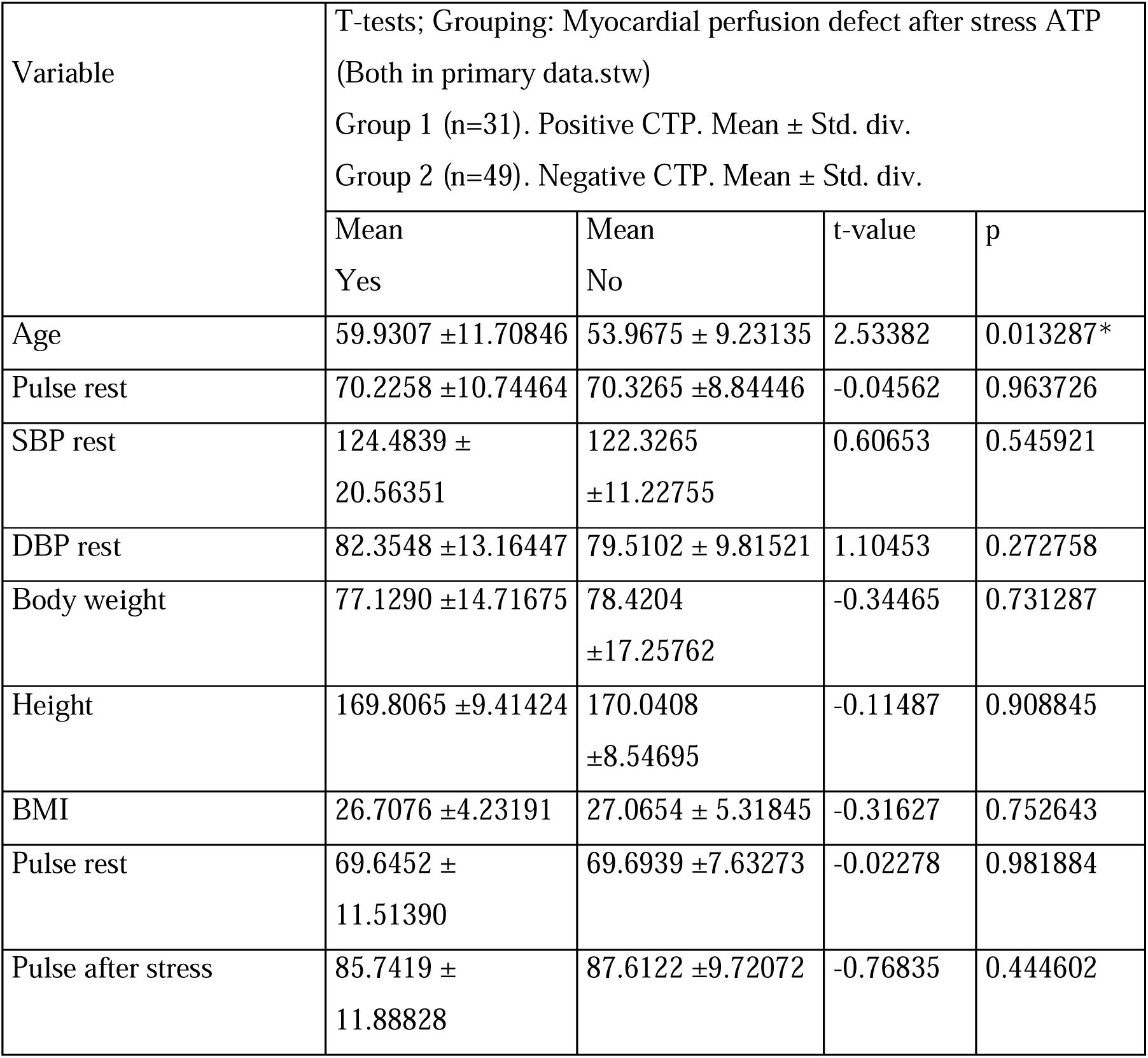
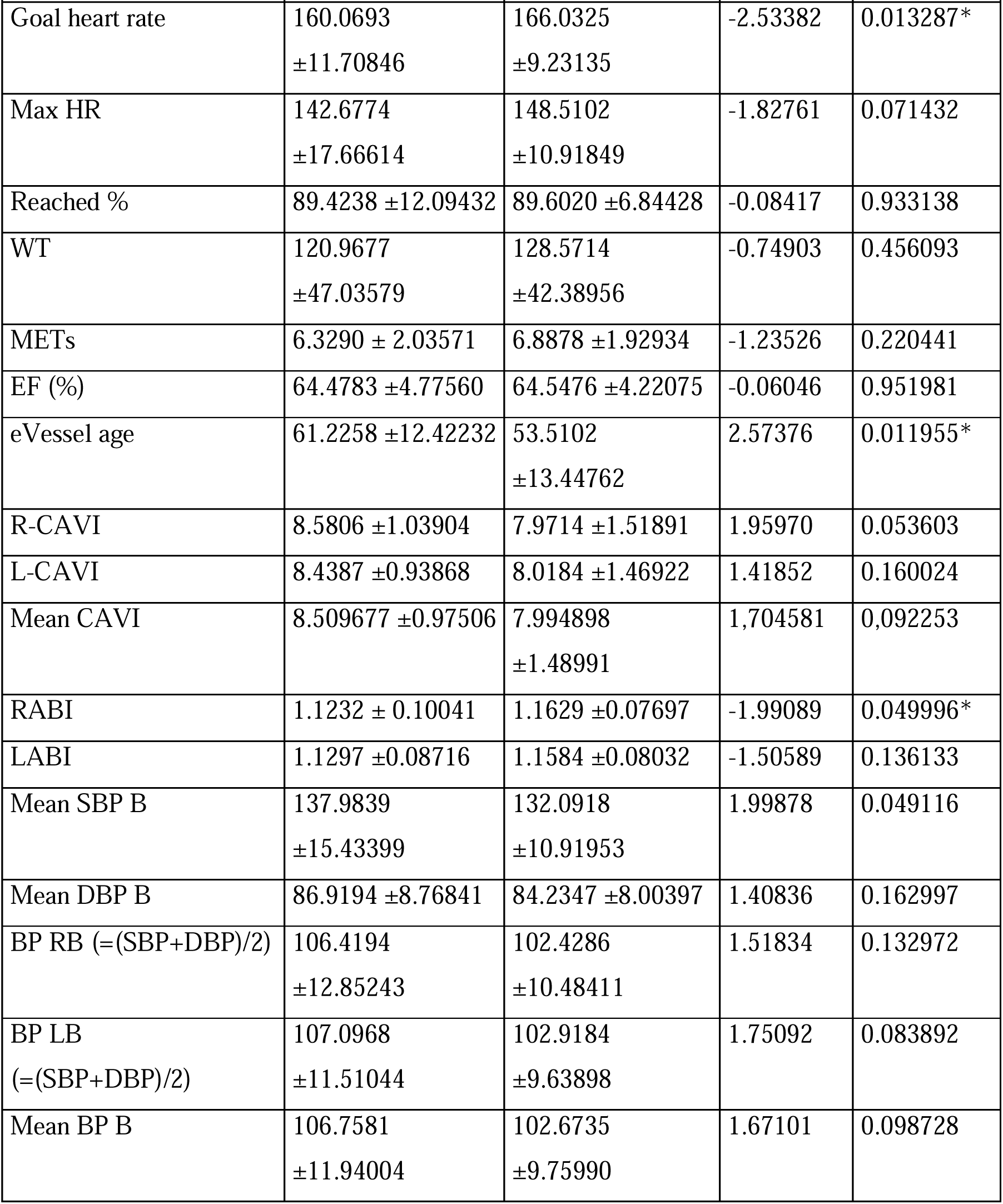
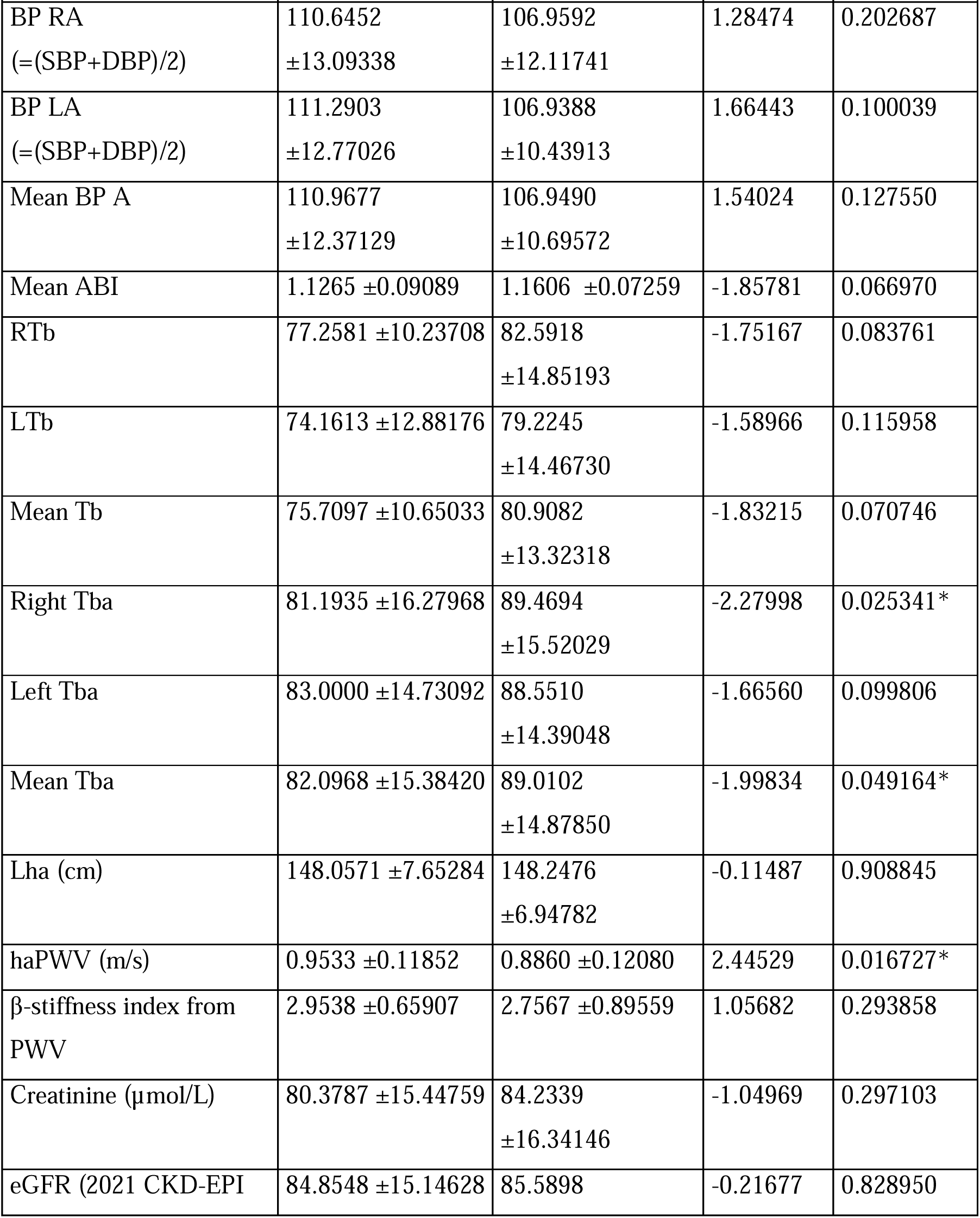
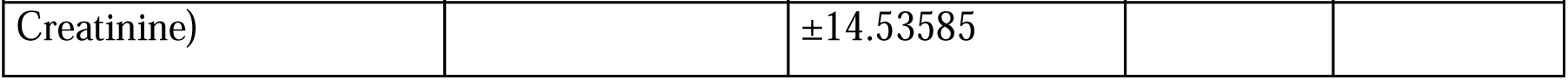
Continuous variables of the sample presented as mean ± standard deviation (Std. div.), Student test as independent variables used. * Values of statically significant differences. Abbreviations: SBP; systolic blood pressure, DBP; diastolic blood pressure, BMI, body mass index, HR; heart rate, METs; metabolic equivalent, R-CAVI; right cardio-ankle vascular index, L-CAVI; left Cardio-ankle vascular index, RABI; right ankle-brachial index, LABI; left ankle-brachial index, SBP B; systolic blood pressure brachial, DBP B; diastolic blood pressure brachial, BP RB; blood pressure right brachial, BP RA; blood pressure right ankle, BP LA; blood pressure left ankle, BP A; blood pressure ankle, ABI; ankle-brachial index, RTb; right brachial pulse, LTb; left brachial pulse, Tb; mean brachial pulse, Tba; mean brachial-ankle pulse, Lha (cm); length heart-ankle, haPWV (m/s); heart-ankle pulse wave velocity.

### The diagnostic accuracy of the bicycle ergometry

We examined the diagnostic accuracy of a standard exercise test on a bicycle ergometer. In the ROC analysis, where the predictor was the result of a sample with the results of the physical exertion “Reaction_type” = ‘Positive’, and the target variable was Myocardial_perfusion_defect_after_stress_ATP, the following results were obtained. (*Table 3*)

**Table 3:**
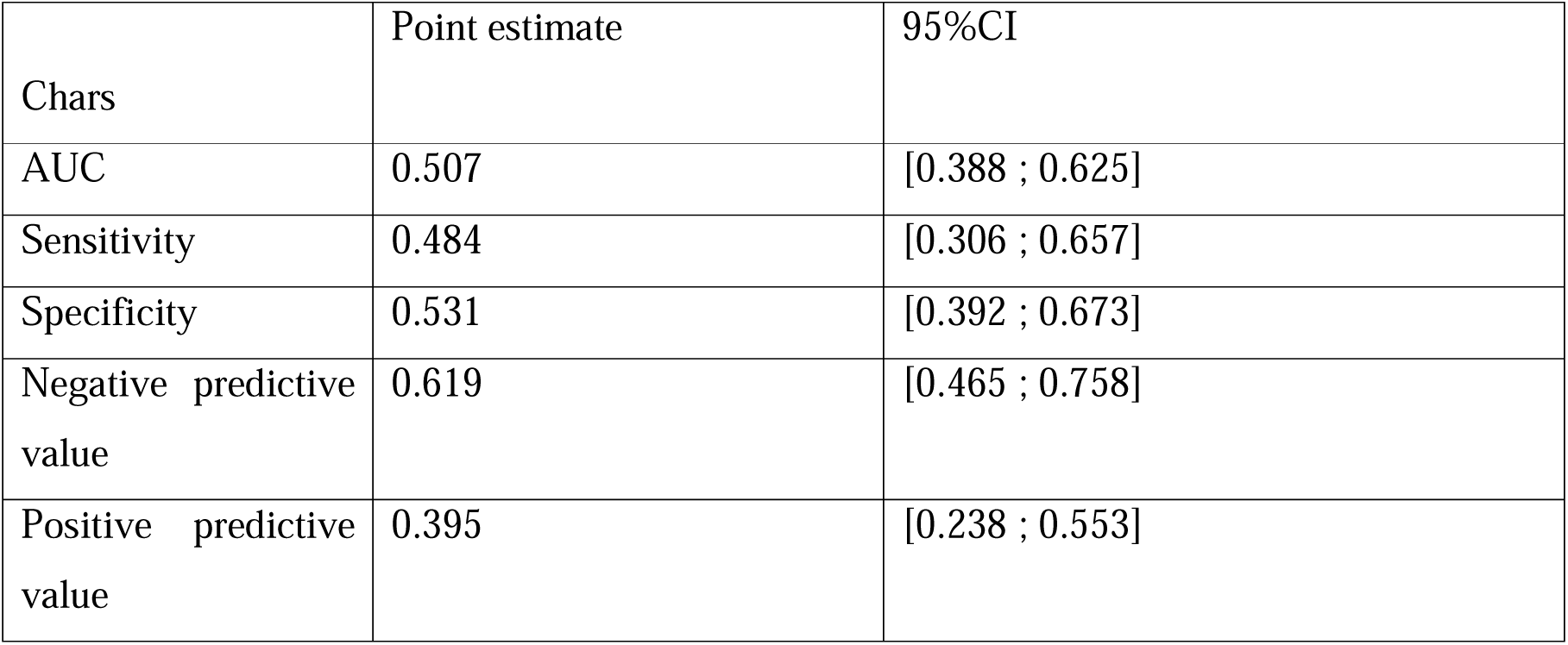
The quality of the bicycle ergometry appeared quite low in our cohort.

### CARDO-QVARK Feature selection with cross-validation

After performing pipeline for feature selection using 1000 iterations, at each iteration, a classifier was built using the gradient boosting algorithm, which made it possible to calculate feature importances for 1000 times. Then, feature importances medians were calculated for each factor, and predictors were ranked from the highest median values to the lowest. The following predictors were selected for Quark, the top 10 based on the median feature importances are presented below. (*Table 4*)

**Table 4:** The 10 most statically significant features according to the build model represented in the table. Dltq_01 indicates the difference between the selected single channel ECG features immediately after the stress test (bicycle ergometry) minus the selected features before the stress test.

|  | Feature | Lasso absolute coefficient |
| --- | --- | --- |
| 1. | q0_QTc | 0.0221996 |
| 2. | q0_Beta | 0.0211857 |
| 3. | q0_J80A | 0.0179913 |
| 4. | q0_Pan...30 | 0.0168176 |
| 5. | q0_SA | 0.013914 |
| 6. | dltq_01_SA | 0.0126805 |
| 7. | dltq_01_Sbeta | 0.0125996 |
| 8. | q0_TA | 0.0124491 |
| 9. | dltq_01_RA | 0.0116579 |
| 10. | q0_Sbeta | 0.0111864 |

The model was then rebuilt as follows. The top 5 predictors from *Table 4* were with the most mathematically importance according to the built model taken and included in the new LASSO regression model.

Then the leave-one-out cross-validation procedure was performed, which allowed us to obtain approximate estimates of sensitivity, specificity, positive and negative prognostic value. At each iteration of leave-one-out cross-validation, the quantitative predictors were normalized.

The quality of the classification is shown in the table below. (*Table 5*)

**Table 5:**
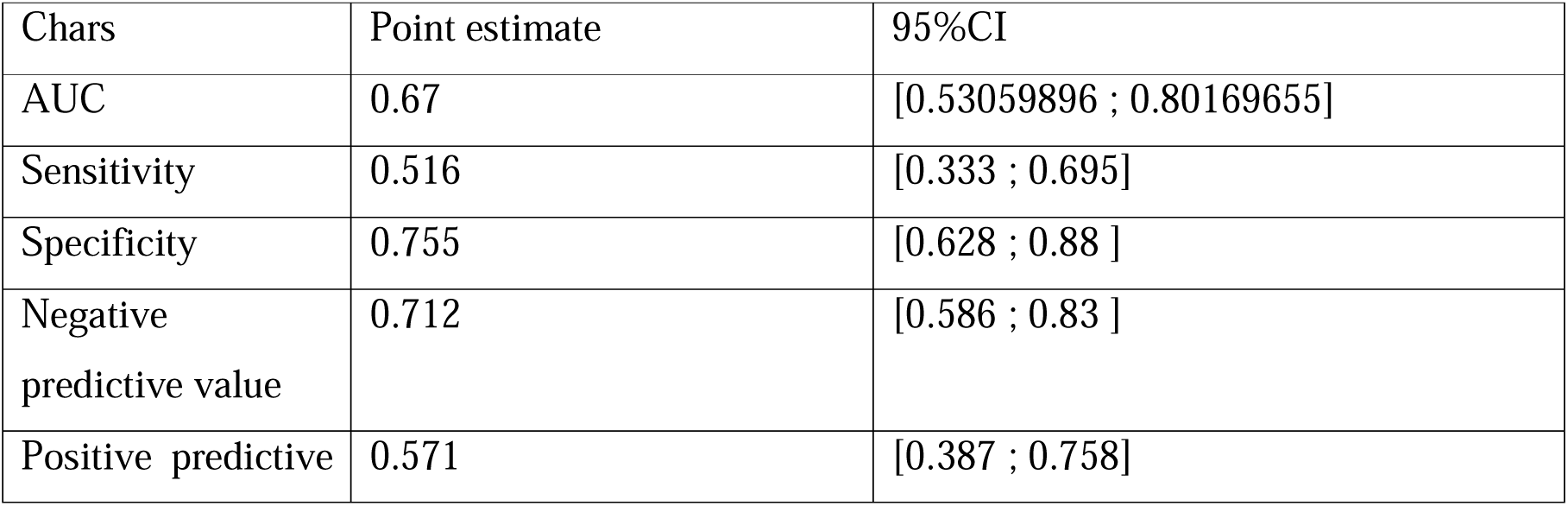
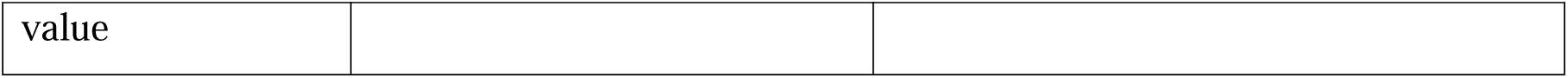
The quality of the single channel ECG (CARDIO-QVARK) in the diagnosis of ischemic heart disease.

Confidence results are calculated using a bootstrap. Comparison with load results was carried out using the McNemar test [37]. (*Figure 1*)

**Figure 1:**
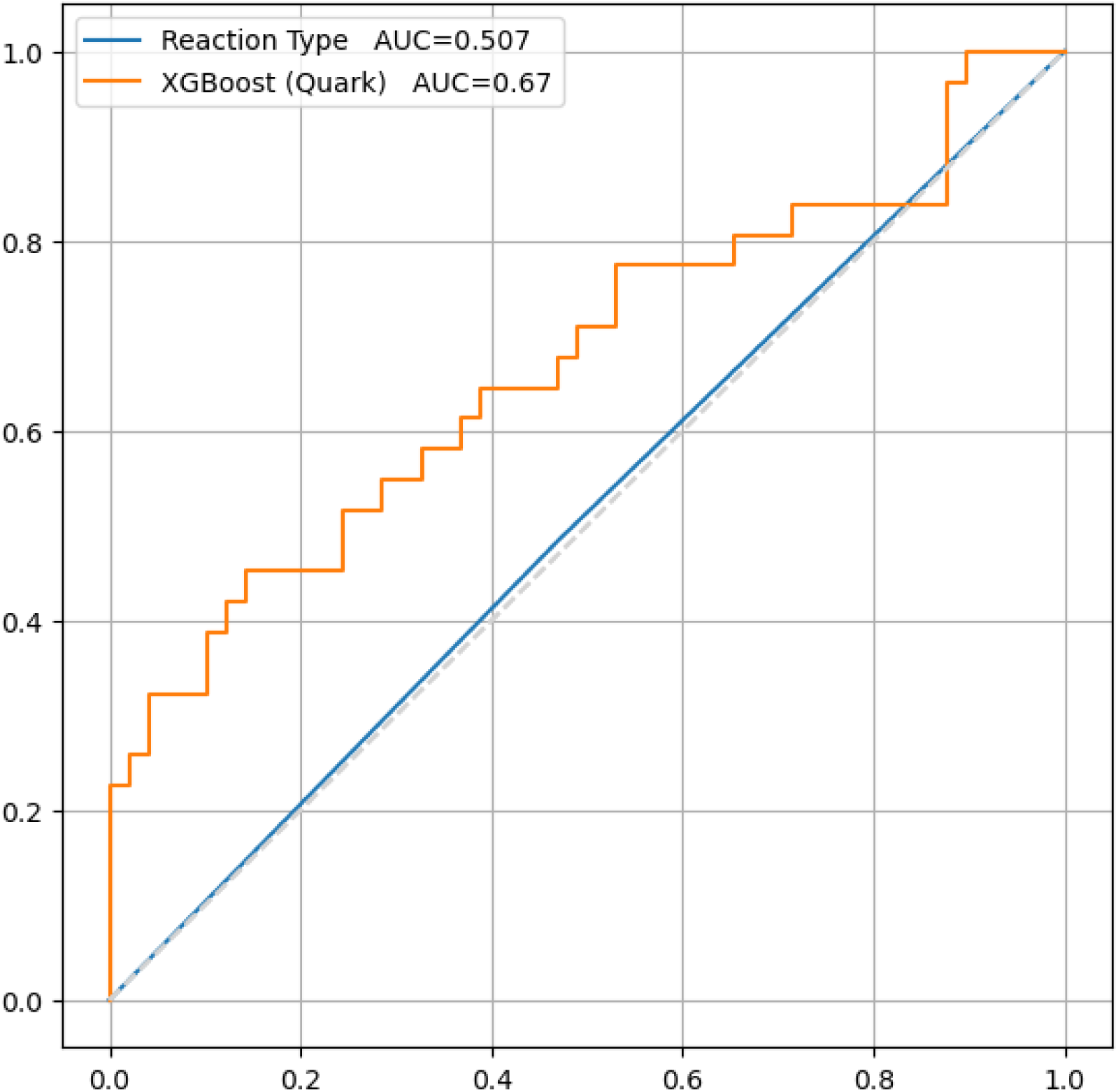
There is no statistically significant difference between the results of the diagnostic accuracy of the load test (50.7 %) and the built model (67.0%), based on our study results. Obviously, the model has better predictive properties, P value = 0.337.

## Discussion

Using single channel ECG in the diagnosis of ischemic heart disease is a potential novel diagnostic strategy that requires further elaboration. Moreover, the usage of single channel ECG in optimizing the physical stress test such as bicycle ergometry is an optimistic strategy [1]. Single channel ECG results can be interpreted using machine learning models to increase the diagnostic accuracy and reduce the consumed time for interpretation of the results by the physicians. Additionally, the primary data of the single channel ECG (approximately 200 parameter) can be used as a novel risk scoring for future IHD development or death due to [10].

Using the physical stress tested monitored 12 lead-ECG remains the elementary test for the primary detection of ischemic heart disease. However, severe limitations exist in the diagnostic accuracy related to the ECG artifacts during the movement of the patients during the physical stress test. Overcoming this issue can be by combining the single channel ECG with the physical stress test, to be performed after the physical stress test immediately and looking for the suggested parameters by our model including the q0_QTc, q0_Beta, q0_J80A, q0_Pan…30, and q0_SA.

Improving the diagnostic accuracy of the physical stress test is a point of focus of the cardiological scientific community. Several attempts performed to enhance the diagnosis performance of the physical exertion tests using complementary methods such as the dynamics of cardiac electrical activity (EAS) during exercise testing [38]. The study suggests that incorporating the equivalent electric cardiac generator of dipole type during exercise ECG testing can enhance the accuracy of diagnosing coronary artery disease [38].

Previous clinical study using a Wearable wireless electrocardiographic (ECG) has shown that the single channel ECG has a poor sensitivity 8.3% (1.0–27.0%) and quite high specificity 89.9% (80.2–95.8%) for detection of reversible ischemic heart disease [39]. The study concluded that both 12-lead ECG (sensitivity 12.5% (3.0–34.4%), specificity 91.3% (82.0– 96.7%)) and the single channel ECG have poor clinical usefulness in terms of ability to detect ischemic heart disease. Interestingly, a dramatic difference has been observed in the II lead of the 12-lead ECG in compare to the single channel ECG [39]. However, other studies suggested the use of deep learning models to enhance the diagnostic accuracy (sensitivity) of the ECG for ischemic heart disease in emergency department [40].

Advancements in single-channel ECG technology for the detection of ischemic heart disease demonstrated that machine learning models based on single-lead ECG and pulse wave parameters, along with age and gender, can simplify screening diagnostics of ejection fraction decrease and diastolic dysfunction with high accuracy [41]. Furthermore, high-frequency ECG signals have shown increased sensitivity and early timing in diagnosing cardiac ischemia, and portable high-resolution ECG devices have demonstrated utility in acute emergency settings [42].

Several ongoing clinical trials to assess the reliability of single channel ECG in the diagnosis of ischemic heart disease and arrythmia in both adults and children (NCT05756309, NCT06181799).

## Conclusions

Single channel ECG (CARDO-QVARK) has the potential to be used as an additional method for the enhancement the diagnostic accuracy of ischemic heart disease in combination with the physical stress test such as bicycle ergometry. Further clinical studies required on a larger sample size to validate the usage of the CARDO-QVARK in clinical practice for the diagnosis of ischemic heart disease.

The following CARDO-QVARK parameter are of interest for further investigation to reveal the hidden diagnostic value in the diagnosis of ischemic heart disease, q0_QTc, q0_Beta, q0_J80A, q0_Pan…30, and q0_SA.

## Data Availability

All data produced in the present work are contained in the manuscript

## List of abbreviations

CVD: cardiovascular disease
CTP: stress computed tomography myocardial perfusion imaging

## Decelerations

1. Ethics approval and consent to participate: the study approved by the Sechenov University, Russia, from “Ethics Committee Requirement № 19-23 from 26.10.2023”. A written consent is taken from the study participants
2. Consent for publication: applicable on reasonable request
3. Availability of data and materials: applicable on reasonable request
4. Competing interests: The authors declare that they have no competing interests regarding publication.
5. Funding’s: The work of Philipp Kopylov and Alexander Suvorov was financed by the government assignment 1023022600020-6 «Application of mass spectrometry and exhaled air emission spectrometry for cardiovascular risk stratification». The work of Basheer Marzoog and Peter Chomakhidze was financed by the Ministry of Science and Higher Education of the Russian Federation within the framework of state support for the creation and development of World-Class Research Center ‘Digital biodesign and personalized healthcare’ № 075-15-2022-304.
6. Authors’ contributions: MB is the writer, researcher, collected and analyzed data, interpreted the results, and revised the final version of the manuscript, AS biostatistical analysis of the sample, PCh revised the paper, and PhK revised the final version of the manuscript. All authors have read and approved the manuscript.
7. Acknowledgments: not applicable
8. Authors’ information: **Basheer Abdullah Marzoog**, World-Class Research Center «Digital Biodesign and Personalized Healthcare», I.M. Sechenov First Moscow State Medical University (Sechenov University), 119991 Moscow, Russia; postal address: Russia, Moscow, 8-2 Trubetskaya street, 119991. (, +79969602820). Scopus ID: 57486338800. **Peter Chomakhidze**, World-Class Research Center «Digital Biodesign and Personalized Healthcare», I.M. Sechenov First Moscow State Medical University (Sechenov University), 119991 Moscow, Russia; postal address: Russia, Moscow, 8-2 Trubetskaya street, 119991. **Alexander Suvorov,** World-Class Research Center «Digital Biodesign and Personalized Healthcare», I.M. Sechenov First Moscow State Medical University (Sechenov University), 119991 Moscow, Russia; postal address: Russia, Moscow, 8-2 Trubetskaya street, 119991. **Philipp Kopylov,** director of the institute of the Research Center «Digital Biodesign and Personalized Healthcare», I.M. Sechenov First Moscow State Medical University (Sechenov University), 119991 Moscow, Russia; postal address: Russia, Moscow, 8-2 Trubetskaya street, 119991. Scopus ID: 6507736224.
9. The paper has not been submitted elsewhere

## STANDARDS OF REPORTING

STROBE guideline has been followed.

